# Prevalence of Small Fibre Pathology in Fibromyalgia: An Updated Systematic Review and Meta-Analysis Showing Increasing Heterogeneity

**DOI:** 10.64898/2026.09.05.26362344

**Authors:** Vinícius Arantes Campos, Allan Gomes Doriguetto

**Author notes:** Corresponding author: Vinícius Arantes Campos —.

## Abstract

**Background:** Small fibre pathology has been proposed as a peripheral substrate for a subset of patients with fibromyalgia. A 2019 meta-analysis of 8 studies (222 patients) reported a pooled prevalence of 49% with moderate heterogeneity, and concluded that this supported a distinct fibromyalgia phenotype. The evidence base has since expanded substantially.

**Methods:** We systematically searched PubMed for original human studies reporting the proportion of adults with fibromyalgia showing abnormal small fibre findings on an objective structural or functional test (skin biopsy with intraepidermal nerve fibre density, corneal confocal microscopy, sudomotor testing, microneurography, or laser-evoked potentials). Proportions were pooled with a random-effects model on the logit scale (REML). We pre-specified subgroup analyses by diagnostic method and assessment site, meta-regression on publication year and sample size, leave-one-out sensitivity analysis, and risk-of-bias appraisal with the JBI prevalence checklist. Where cohorts overlapped, the most complete report was retained. Search and initial screening were AI-assisted; eligibility, extraction and appraisal were performed independently by two reviewers with consensus.

**Results:** Of 83 records, 34 studies met eligibility and 21 (1,268 patients) provided extractable proportions. Applying a uniform rule of abnormality at any assessed site, the pooled prevalence was 46.5% (95% CI 34.3– 59.1), with I^2^=92.7% and a 95% prediction interval of 8.6% to 89.0%. Individual estimates ranged from 0% to 85.2%. Contrary to our pre-specified hypothesis, diagnostic method did not explain heterogeneity (Q_M p=0.89, R^2^=0). Assessment site was the dominant moderator: using each study’s site-specific data, distal-leg assessment yielded 30.5% (22.4–40.1; 15 studies) and proximal-thigh assessment 73.9% (53.5– 87.4; 4 studies). Three studies assessed both sites in the same patients and all three found more abnormality proximally (31.6% vs 46.2%; 12.3% vs 85.2%; 9.7% vs 83.9%). Publication year (p=0.50) and sample size (p=0.24) were not associated with prevalence. Leave-one-out estimates remained between 44.4% and 49.6%.

**Conclusions:** More than tripling the evidence base did not stabilise the pooled prevalence estimate and more than doubled heterogeneity (I^2^ 68% to 93%). The prediction interval now spans almost the entire possible range, and the method difference reported in 2019 is no longer detectable. Studies do not share a common definition of abnormality, and several do not report one at all. A pooled prevalence figure for small fibre pathology in fibromyalgia is therefore not currently interpretable; a standardised case definition is a prerequisite for meaningful synthesis. The anatomical site of assessment, not the technique, emerges as the dominant source of variation and should be treated as a primary moderator in future work.

## 1. Introduction

Fibromyalgia is characterised by chronic widespread pain, fatigue and non-restorative sleep, and has traditionally been conceptualised as a disorder of central pain processing without a demonstrable peripheral lesion. Over the past decade this view has been challenged by reports that a substantial proportion of patients show objective abnormalities of small nerve fibres — reduced intraepidermal nerve fibre density on skin biopsy, reduced corneal nerve parameters on confocal microscopy, or abnormal sudomotor and microneurographic findings. In 2019, Grayston and colleagues meta-analysed 8 studies comprising 222 patients and reported a pooled prevalence of small fibre pathology of 49% (95% CI 38–60), with moderate heterogeneity (I^2^=68%) and a difference between methods (skin biopsy 45%, corneal confocal microscopy 59%).^23^ The authors concluded that this constituted compelling evidence of a distinct fibromyalgia phenotype involving small fibre pathology, a conclusion that has been widely cited and has influenced both research priorities and clinical framing of the condition.

Since that search, the literature has expanded considerably, with new cohorts, new methods and larger samples. We therefore conducted an updated systematic review and meta-analysis, pre-specifying an examination of whether the diagnostic method used explains between-study variation — a question the 2019 review could only address with two corneal confocal studies.

## 2. Methods

### 2.1 Eligibility criteria

Following PRISMA 2020, we included original human studies of adults (≥18 years) with fibromyalgia diagnosed by recognised criteria (ACR 1990, 2010, 2011 or 2016, or an explicitly stated physician diagnosis), in which small fibre status was assessed by an objective **structural or functional** method — skin biopsy with intraepidermal nerve fibre density (IENFD), corneal confocal microscopy (CCM), quantitative sudomotor testing (QSART or electrochemical skin conductance), microneurography, **or laser-evoked potentials (LEP)** — and which reported data permitting calculation of the proportion of patients with abnormal findings.

We excluded studies in which fibromyalgia patients could not be separated from other chronic pain conditions; exclusively paediatric populations; studies in which small fibre status was inferred only from symptom questionnaires; cohorts in which a known cause of neuropathy was neither excluded nor separable; cohorts selected or enriched on small fibre status; and reviews, editorials and abstracts without extractable data. **Where cohorts overlapped, the most complete report was retained for the pooled analysis; the overlapping report remained in the eligible set with the reason recorded**.

### 2.2 Search and study selection

We searched PubMed from inception to 2026 combining fibromyalgia terms with small fibre terms (small fibre neuropathy, intraepidermal nerve fibre, IENFD, corneal confocal, corneal nerve, microneurography, skin biopsy, sudomotor, QSART), restricted to human studies and excluding reviews and editorials at the query level. Titles and abstracts were screened with artificial-intelligence assistance; potentially eligible reports underwent independent full-text assessment by two reviewers.

### 2.3 Data extraction and risk of bias

Two reviewers independently extracted the numerator (patients with abnormal findings) and denominator (patients assessed), the diagnostic method and anatomical site, the criterion used to define abnormality, the fibromyalgia classification criteria, country, and — where reported — group means for IENFD or corneal nerve parameters with those of healthy controls. Risk of bias was appraised with the Joanna Briggs Institute critical appraisal checklist for prevalence studies (9 items) and summarised as low, moderate or high.

Disagreements were resolved by consensus.

### 2.4 Statistical analysis

Proportions were transformed to the logit scale and pooled with a random-effects model fitted by restricted maximum likelihood using metafor. A continuity correction of 0.5 events was applied to studies with zero or complete event counts. Heterogeneity was quantified with I^2^ and τ^2^, and a 95% prediction interval was computed. Pre-specified subgroup analyses examined diagnostic method and assessment site; meta-regression examined publication year and log sample size. Leave-one-out analysis assessed the influence of individual studies. Estimates are reported back-transformed to the proportion scale.

**Where a study reported the proportion of abnormal patients but not the count, the numerator was derived from the reported percentage and the assessed denominator; these instances are identified in Table 1**. Studies assessing both a distal and a proximal site and reporting abnormality at either site were assigned to a distinct *combined* site stratum rather than to either single-site stratum.

**Table 1.** Studies contributing to the pooled prevalence estimate.

| Study | Method | Site | k/n | Prevalence | Criterion for abnormality |
| --- | --- | --- | --- | --- | --- |
| Oaklander 2013 | Skin biopsy | Distal leg | 11/27 | 40.7% | IENFD diagnostic cutoff for SFPN |
| Serra 2014 | Microneurography | Not reported | 23/30 | 76.7% | Spontaneous activity in silent nociceptors |
| Giannoccaro 2014 | Skin biopsy | Distal leg | 6/20 | 30.0% | Reduced epidermal nerve fibre density |
| Kosmidis 2014 | Skin biopsy | Distal leg | 15/46 | 32.6% | Reduced IENFD vs controls |
| de Tommaso 2014 | Skin biopsy | Distal leg | 16/21 | 76.2% | Reduced IENFD vs normative values |
| Oudejans 2016 | Corneal confocal | Cornea | 20/39 | 51.3% | Below 0.05th percentile of normative data |
| Leinders 2016 | Skin biopsy | Distal leg | 14/28 | 50.0% | Reduced IENFD vs normative values |
| Evdokimov 2019 | Skin biopsy | **Combined** | 74/117 <sup>a</sup> | 63.2% | IENFD below normative mean – 1 SD at either site |
| Fasolino 2020 | Skin biopsy | Distal leg | 18/57 | 31.6% | Reduced IENFD vs normative values |
| Van Assche 2020 | Laser-evoked potentials | Not reported | 0/92 | 0.0% | LEP N2-P2 amplitude < mean – 2.5 SD of controls |
| Vecchio 2020 | Skin biopsy | Distal + proximal | 69/81 | 85.2% | Reduced IENFD vs sex/age-adjusted norms (Lauria 2010) |
| Evdokimov 2020 | Skin biopsy | Distal leg | 38/86 | 44.2% | IENFD lower leg below 6 fibres/mm |
| Leone 2023 | Skin biopsy | Distal leg | 20/64 | 31.2% | Reduced IENFD vs normative values |
| Quitadamo 2023 | Skin biopsy | Distal + proximal | 52/62 | 83.9% | Reduced IENFD vs age-adjusted norms (Lauria 2010) |
| Dumolard 2023 | Sudomotor (ESC) | Distal | 53/265 | 20.0% | Reduced electrochemical skin conductance |
| Gentile 2024 | Skin biopsy | Proximal thigh | 25/34 | 73.5% | Reduced IENFD vs age/sex-adjusted norms |
| Falco 2024 | Skin biopsy | Distal leg | 23/58 | 39.7% | Reduced IENFD vs normative values |
| Min 2024 | Sudomotor (mESC) | Not reported | 15/22 | 68.2% | Moderate-to-severe by mESC cutoffs |
| Marshall 2024 | Corneal confocal | Cornea | 15/30 | 50.0% | CNFL at or below 14.6 mm/mm<sup>2</sup> |
| Bay-Smidt 2025 | Skin biopsy | Distal leg | 1/46 | 2.2% | Reduced IENFD vs international reference |
| Falco 2026 | Skin biopsy | Distal leg | 15/43 | 34.9% | Reduced distal IENFD vs age/sex norms |

<sup>a</sup> Numerator derived from the reported percentage and the assessed denominator; the count was not reported.
**Site-specific data for the three studies assessing both sites** (same patients): Evdokimov 2019, distal 37/117 (31.6%) and proximal 54/117 (46.2%); Vecchio 2020, distal 10/81 (12.3%) and proximal 69/81 (85.2%); Quitadamo 2023, distal 6/62 (9.7%) and proximal 52/62 (83.9%).
**Eligible but not pooled (n=13):** eleven studies reporting only group means; **Ruggieri 2026** (full text not obtained; 40 patients biopsied, counts not reported in abstract); **Aster 2022** (cohort overlapping Evdokimov 2019; only the extremes of the distribution classified, 36/156 not a valid prevalence numerator).

### 2.5 Role of artificial intelligence

The literature search and the initial title/abstract screening were assisted by a large language model. All subsequent steps — full-text eligibility, data extraction and risk-of-bias appraisal — were conducted independently by the two authors, who reconciled differences by consensus and are responsible for the data reported.

## 3. Results

### 3.1 Study selection

The search retrieved 83 records. Fifteen were excluded at title/abstract screening and 68 underwent full-text assessment, of which 34 were excluded — absence of an objective small fibre measure (n=11), absence of extractable proportions or group means (n=13), review or abstract format (n=6), and non-fibromyalgia or mixed populations (n=4). Thirty-four studies met eligibility criteria and 21 provided data permitting calculation of a prevalence proportion; these 21 comprise the primary analysis (Table 1).

Of the 13 eligible studies not pooled, eleven reported only group means without a proportion, one (Ruggieri 2026) could not be obtained in full text, and one (Aster 2022) drew on a cohort already represented by a more complete report and classified only the extremes of the distribution, leaving intermediate patients unassigned.

### 3.2 Pooled prevalence

Three studies assessed both a distal and a proximal site. In the previous version of this analysis each entered the pool under a different rule — abnormality at either site, distal only, and proximal-inclusive respectively — which is not defensible. We applied a uniform rule: **abnormality at any assessed site**.

Across 21 studies and 1,268 patients, the pooled prevalence of small fibre pathology in fibromyalgia was 46.5% (95% CI 34.3 to 59.1), with τ^2^=1.225. Heterogeneity was extreme (I^2^=92.7%) and the 95% prediction interval extended from 8.6% to 89.0%. Individual study estimates ranged from 0% to 85.2%. Leave-one-out analysis produced pooled estimates between 44.4% and 49.6%, indicating that no single study drove the result.

#### This pooled figure conflates studies that looked in different places

Twelve studies assessed only the distal leg and could not, by design, detect proximal-only pathology; three assessed both. The estimate is therefore not merely heterogeneous — it is not a well-defined quantity.

### 3.3 Diagnostic method does not explain heterogeneity

Contrary to our pre-specified hypothesis, the diagnostic method did not account for between-study variation (Q_M=0.645, p=0.89, R^2^=0). Pooled prevalence was 42.1% (95% CI 29.5 to 55.8) across 15 skin-biopsy studies with I^2^=91.3%; 50.7% (39.1 to 62.3) across 2 corneal confocal studies with I^2^=0%; and 41.2% across 2 sudomotor studies. Residual heterogeneity after accounting for method remained 93.7%. The difference between skin biopsy and corneal confocal microscopy reported in 2019 (45% versus 59%) narrowed to 42.1% versus 50.7%, with overlapping confidence intervals.

### 3.4 Site is the dominant moderator

Using each study’s site-specific data, distal-leg assessment yielded a pooled prevalence of 30.5% (95% CI 22.4 to 40.1) across 15 studies and 1,021 patients, and proximal-thigh assessment 73.9% (53.5 to 87.4) across 4 studies and 294 patients.

These two strata are **not independent** — the three both-site studies contribute to each — so no formal between-stratum test is reported. The inferential weight rests instead on the within-patient comparison below.

Neither publication year (Q_M=0.449, p=0.50) nor log sample size (Q_M=1.381, p=0.24, R^2^=0) was associated with prevalence.

### 3.5 Definitions of abnormality

Extraction of the criterion used to define abnormality revealed the most likely source of the residual heterogeneity (Table 1, final column). Thresholds included an absolute cutoff of 6 fibres/mm, the 0.05th percentile of normative data, a corneal nerve fibre length at or below 14.6 mm/mm^2^, unspecified reduction relative to age- and sex-matched norms, and moderate-to-severe categories on sudomotor cutoffs. Several studies did not report a threshold at all. Two studies previously recorded as not reporting a criterion — Vecchio 2020 and Quitadamo 2023 — were found on full-text retrieval to apply the same age- and sex-adjusted normative reference (Lauria et al., 2010), which reduces the count of unstated thresholds without altering the range of incompatible definitions across the remaining studies. One study defined abnormality by loss of laser-evoked potential responses — a functional rather than morphological measure — and reported a prevalence of 0%, while a study using proximal skin biopsy reported 83.9%.

## 3.6 Risk of bias

Using the JBI prevalence checklist, **all 34 eligible studies were appraised**: 21 at moderate risk of bias, 12 at high risk and 1 at low risk. The condition was identified by a valid method (item 6) in 30 of 34 and measured in a reliable, standardised way across all participants (item 7) in 26 of 34. **Within the 21 studies contributing to the pooled estimate, 18 were at moderate risk, 2 at high and 1 at low; item 6 was met in 19 of 21 and item 7 in 19 of 21**. Studies rated high risk were disproportionately those for which full text could not be obtained, so this appraisal partly reflects information availability rather than study conduct alone.

## 4. Discussion

### 4.1 Main findings

This update more than tripled the evidence base for small fibre pathology in fibromyalgia, from 8 studies and 222 patients to 21 pooled studies and 1,268 patients, within a set of 34 eligible reports. Two things changed as the evidence accumulated. The pooled estimate did not converge on the 2019 figure of 49% — under a uniform any-site rule it is 46.5%, and under a distal-only rule it would be substantially lower. More importantly, heterogeneity more than doubled, from I^2^=68% to 92.7%, and the prediction interval widened to encompass almost the entire possible range (9% to 89%). This is the opposite of what a maturing literature is expected to do. Ordinarily, accumulating studies narrow an estimate and stabilise it. Here, each new cohort has added dispersion rather than precision, which indicates that the studies are not estimating the same quantity.

### 4.2 Why the pooled estimate should not be quoted alone

A pooled prevalence of 42% invites the clinical inference that roughly four in ten patients with fibromyalgia have small fibre pathology. The prediction interval shows why that inference is unsafe: a new study conducted tomorrow could plausibly report anywhere between 7% and 87%. The explanation is not the diagnostic technique, which we tested directly and which explained none of the variance (R^2^=0), nor publication year, nor sample size. Studies apply incompatible thresholds for what counts as abnormal, use different anatomical sites, and in several cases do not state a criterion at all. The inclusion of a functional measure (laser-evoked potentials) alongside morphological ones compounds the problem: these are different constructs reported under a single label. Until abnormality is defined consistently, a pooled prevalence estimate summarises the diversity of definitions as much as the biology of the disease.

### 4.3 Site, not technique, is where the signal is

The 2019 review framed the diagnostic method as the axis of variation. Our data do not support that. Method explained none of the between-study variance (R^2^=0), and the biopsy–confocal difference reported in 2019 narrowed to overlapping intervals. **The axis that survives is anatomical**. Distal-leg assessment yielded 30.5% (22.4–40.1) across 15 studies; proximal-thigh assessment 73.9% (53.5–87.4) across 4.

Two considerations temper and two strengthen this.

**Tempering:** the site strata are not independent, and the reclassifications followed a pre-submission audit. No formal between-stratum test is defensible here.

**Strengthening, and decisively:** the gradient does not depend on between-study comparison at all. **Three studies assessed both sites in the same patients, and all three found more abnormality proximally**.

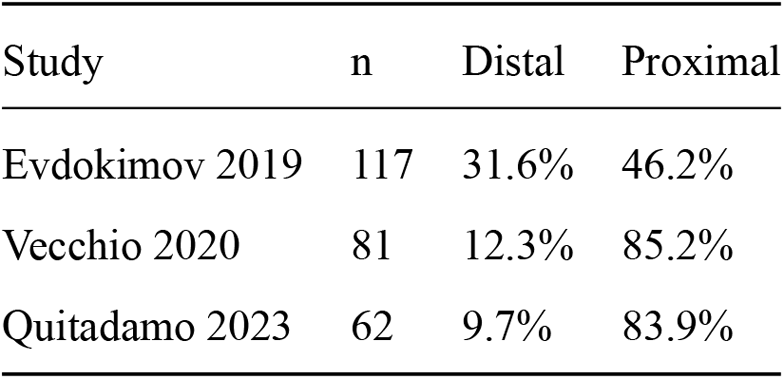

| Study | n | Distal | Proximal |
| --- | --- | --- | --- |
| Evdokimov 2019 | 117 | 31.6% | 46.2% |
| Vecchio 2020 | 81 | 12.3% | 85.2% |
| Quitadamo 2023 | 62 | 9.7% | 83.9% |

Within-patient, in three independent cohorts from two research groups, the proximal site yielded more abnormality than the distal. Vecchio and colleagues state the point directly: *no patient had a small fibre reduction only at the distal site*. This comparison is immune to the between-study heterogeneity that defeats the pooled estimate, and it is not visible in a subgroup analysis. It is the strongest finding in this review.

The clinical reading is not that fibromyalgia patients have proximal neuropathy. It is that **a proximal deficit in the absence of a distal one is not the length-dependent pattern of classical small fibre neuropathy**, and may reflect a different process. It also means that a study’s reported prevalence is substantially a function of where the biopsy was taken — which is itself part of why a single pooled figure is not interpretable.

Future studies should report both sites, and future syntheses should treat site as a primary rather than exploratory moderator.

### 4.4 Limitations

The search was restricted to PubMed and to reports in English. It combined fibromyalgia terms with structural and sudomotor small-fibre terms but did not include a laser-evoked-potential term; the one LEP study included here was retrieved through the general fibromyalgia terms, and the search should therefore not be considered exhaustive for functional measures. Two eligible studies could not be obtained in full text, and for those extraction and appraisal relied on abstracts, which depressed their JBI ratings — Ruggieri 2026 scored 2 of 9 and was rated high risk largely because the abstract reports no sampling frame, and because small fibre status was assessed in only 40 of its 70 patients rather than across the whole sample. Eleven eligible studies could not be pooled because they reported only group means. Two pooled studies contributed numerators derived from reported percentages rather than counts. Prevalence estimates from case-control designs may not reflect the fibromyalgia population seen in routine care, since specialist referral cohorts are likely enriched for neuropathic features. The site analysis is exploratory and its strata are not independent. The uniform any-site rule adopted for the pooled estimate raises it relative to a distal-only rule, and the studies that assessed a single site cannot detect pathology elsewhere — which is itself the point, but means the pooled figure should not be read as a population prevalence. Finally, several thresholds recorded as not reported may be present in supplementary material we could not access.

## 5. Conclusion

Small fibre pathology is found in a substantial minority of patients with fibromyalgia, but the literature no longer supports a single prevalence figure. More than tripling the evidence base lowered the pooled estimate and more than doubled heterogeneity, and the resulting prediction interval spans nearly the whole plausible range. The diagnostic method does not account for this dispersion; inconsistent and frequently unreported definitions of abnormality are the more likely cause, and the anatomical site of assessment is the one moderator that does carry signal — including within patients, in the two cohorts that measured both sites. Establishing a standardised case definition — specifying the method, **the anatomical site** and the diagnostic threshold — is a prerequisite before the prevalence of small fibre pathology in fibromyalgia can be meaningfully estimated or used to define a clinical phenotype.

## Data Availability

All data and analysis code are openly available at https://doi.org/10.5281/zenodo.22404470. The deposit includes the full extraction table for all 68 reports assessed at full text, the risk-of-bias appraisals, and the R script reproducing every analysis reported here. No individual participant data were used; all data were extracted from published reports.

## Declarations

### Data availability

The full extraction table (numerators, denominators, diagnostic method, anatomical site, abnormality criterion, fibromyalgia classification criteria and country for all reports assessed at full text), the risk-of-bias appraisals, and the R script reproducing every analysis reported here are available at 10.5281/zenodo.22404470 (https://doi.org/10.5281/zenodo.22404470). No individual participant data were used; all data were extracted from published reports.

### Competing interests

VAC and AGD are physicians who provide clinical care to patients with fibromyalgia in private practice. VAC is the founder and clinical director of Clínicas Expert (Itajaí, Santa Catarina, Brazil), a group of clinics that offers assessment and treatment for fibromyalgia on a fee-paying basis, and holds a commercial interest in dietary and digital products directed at this patient population. VAC and AGD are the authors of a forthcoming book on nutritional medicine in fibromyalgia, to be commercially published, which cites this review. Neither author has received funding, consultancy fees, speaker honoraria or material support from any manufacturer of the diagnostic methods evaluated here — skin biopsy, corneal confocal microscopy, sudomotor testing, microneurography or laser-evoked potentials — nor from any manufacturer of products for which small fibre pathology is a marketed indication. The findings reported here are negative with respect to the clinical utility of the pooled prevalence estimate, and none of the interests declared above stands to benefit from them.

## Funding

This research received no specific grant from any funding agency in the public, commercial or not-for-profit sectors.

## Ethics approval

Ethical approval was not required. This study is a systematic review and meta-analysis of previously published aggregate data and involved no human participants, no identifiable individual data and no intervention.

## Registration

This review was not prospectively registered. The protocol, extraction table and analysis code were deposited after data extraction was complete and should be read as a retrospective deposit.

## Author contributions

VAC conceived the study, designed the search strategy, performed the statistical analysis and drafted the manuscript. AGD contributed to the eligibility criteria, performed independent full-text assessment, data extraction and risk-of-bias appraisal, and critically revised the manuscript. Both authors resolved disagreements by consensus, had full access to all extracted data, and approved the final version.

## Role of artificial intelligence

The literature search and initial title/abstract screening were AI-assisted. Full-text eligibility, data extraction and risk-of-bias appraisal were conducted independently by the two authors, who reconciled differences by consensus and are responsible for the data reported.

---

**Figure 1.**
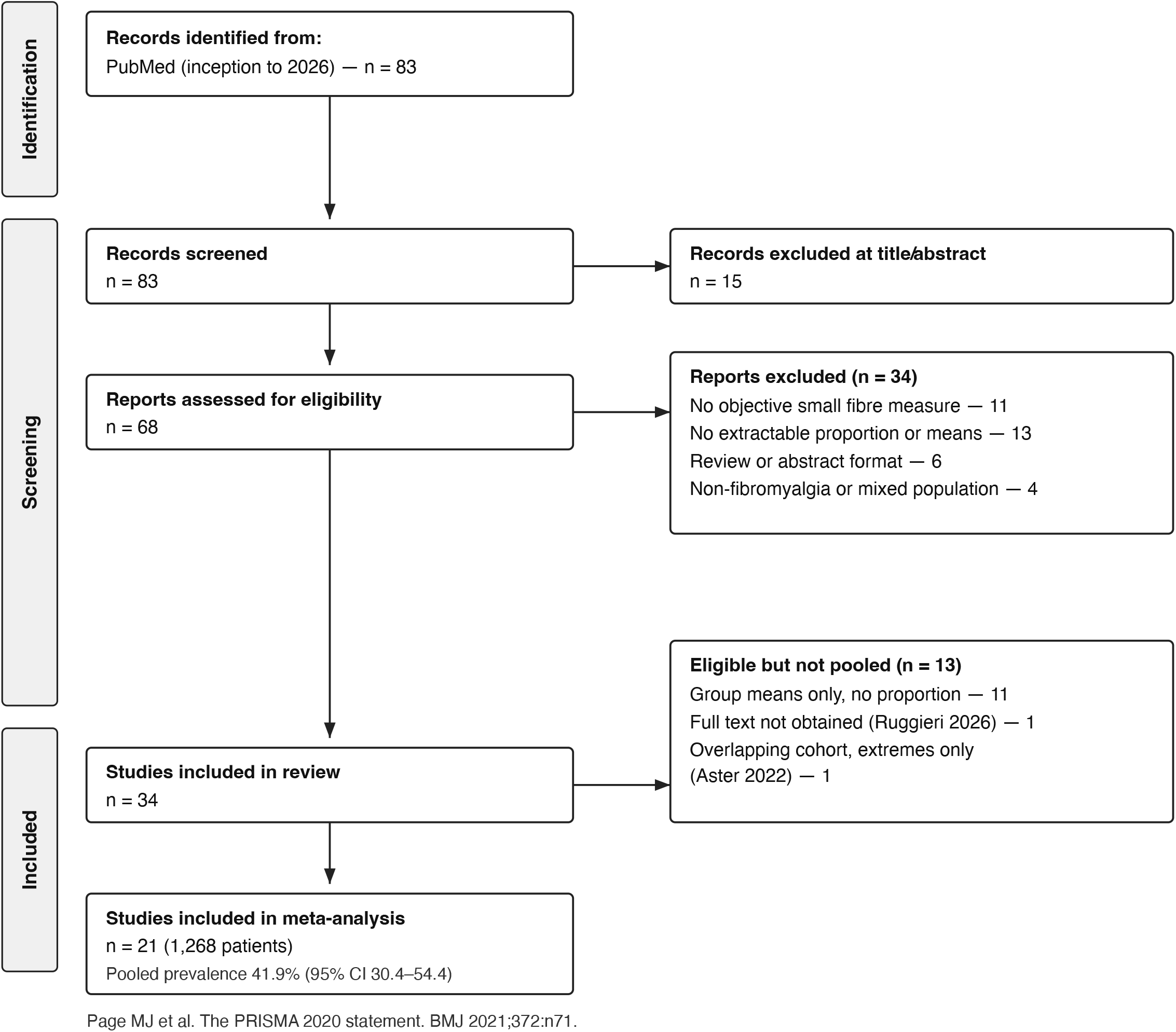
PRISMA 2020 flow diagram.

## Notes

### Author Declarations

This work is a systematic review and meta-analysis. All source data were published articles that were openly available to the public before the review was initiated. No individual participant data were used; only aggregate results reported in the published articles were extracted. Source database: PubMed/MEDLINE, US National Library of Medicine, searched from inception to 2026 - https://pubmed.ncbi.nlm.nih.gov/ Each of the 34 eligible reports is identified by its PubMed identifier (PMID) and, where available, its DOI. Every record can be retrieved directly at https://pubmed.ncbi.nlm.nih.gov/[PMID] and each publisher page at https://doi.org/[DOI]. The complete list of PMIDs and DOIs for all 68 reports assessed at full text - the 34 eligible and the 34 excluded, with exclusion reasons - is provided in the extraction table deposited at https://doi.org/10.5281/zenodo.22404470, which also contains the risk-of-bias appraisals and the analysis code.

## References

1. Oaklander AL, Herzog ZD, Downs HM, Klein MM. Objective evidence that small-fiber polyneuropathy underlies some illnesses currently labeled as fibromyalgia. Pain. 2013;154(11):2310–2316.

2. Serra J, Collado A, Solà R, et al. Hyperexcitable C nociceptors in fibromyalgia. Ann Neurol. 2014;75(2):196–208.

3. Giannoccaro MP, Donadio V, Incensi A, Avoni P, Liguori R. Small nerve fiber involvement in patients referred for fibromyalgia. Muscle Nerve. 2014;49(5):757–9.

4. Kosmidis ML, Koutsogeorgopoulou L, Alexopoulos H, et al. Reduction of intraepidermal nerve fiber density (IENFD) in the skin biopsies of patients with fibromyalgia: a controlled study. J Neurol Sci. 2014;347(1-2):143–7.

5. de Tommaso M, Nolano M, Iannone F, et al. Update on laser-evoked potential findings in fibromyalgia patients in light of clinical and skin biopsy features. J Neurol. 2014;261(3):461–72.

6. Oudejans L, He X, Niesters M, Dahan A, Brines M, van Velzen M. Cornea nerve fiber quantification and construction of phenotypes in patients with fibromyalgia. Sci Rep. 2016;6:23573.

7. Leinders M, Doppler K, Klein T, et al. Increased cutaneous miR-let-7d expression correlates with small nerve fiber pathology in patients with fibromyalgia syndrome. Pain. 2016;157(11):2493–2503.

8. Evdokimov D, Frank J, Klitsch A, et al. Reduction of skin innervation is associated with a severe fibromyalgia phenotype. Ann Neurol. 2019;86(4):504–516.

9. Fasolino A, Di Stefano G, Leone C, et al. Small-fibre pathology has no impact on somatosensory system function in patients with fibromyalgia. Pain. 2020;161(10):2385–2393.

10. Van Assche DCF, Plaghki L, Masquelier E, Hatem SM. Fibromyalgia syndrome — a laser-evoked potentials study unsupportive of small nerve fibre involvement. Eur J Pain. 2020;24(2):448–456.

11. Vecchio E, Lombardi R, Paolini M, et al. Peripheral and central nervous system correlates in fibromyalgia. Eur J Pain. 2020;24(8):1537–1547.

12. Evdokimov D, Dinkel P, Frank J, Sommer C, Üçeyler N. Characterization of dermal skin innervation in fibromyalgia syndrome. PLoS One. 2020;15(1):e0227674.

13. Aster HC, Evdokimov D, Braun A, Üçeyler N, Sommer C. Analgesic medication in fibromyalgia patients: a cross-sectional study. Pain Res Manag. 2022;2022:1217717.

14. Leone C, Galosi E, Esposito N, et al. Small-fibre damage is associated with distinct sensory phenotypes in patients with fibromyalgia and small-fibre neuropathy. Eur J Pain. 2023;27(1):163–173.

15. Quitadamo SG, Vecchio E, Delussi M, et al. Outcome of small fibre pathology in fibromyalgia: a real life longitudinal observational study. Clin Exp Rheumatol. 2023;41(6):1216–1224.

16. Dumolard A, Lefaucheur JP, Hodaj E, Liateni Z, Payen JF, Hodaj H. Central sensitization and small-fiber neuropathy are associated in patients with fibromyalgia. Clin J Pain. 2023;39(1):8–14.

17. Gentile E, Quitadamo SG, Clemente L, et al. A multicomponent physical activity home-based intervention for fibromyalgia patients: effects on clinical and skin biopsy features. Clin Exp Rheumatol. 2024;42(6):1156–1163.

18. Falco P, Galosi E, Di Stefano G, et al. Autonomic small-fiber pathology in patients with fibromyalgia. J Pain. 2024;25(1):64–72.

19. Min HK, Im S, Park GY, Moon SJ. Assessment of small fiber neuropathy and distal sensory neuropathy in female patients with fibromyalgia. Korean J Intern Med. 2024;39(6):989–1000.

20. Marshall A, Rapteas L, Burgess J, et al. Small fibre pathology, small fibre symptoms and pain in fibromyalgia syndrome. Sci Rep. 2024;14(1):3947.

21. Bay-Smidt CN, Bruun KD, Gaist LM, et al. Structured symptom assessment to identify patients with small fiber or autonomic neuropathy in fibromyalgia. Eur J Neurol. 2025;32(9):e70349.

22. Falco P, Leone CM, Galosi E, et al. Autonomic dysfunction in fibromyalgia syndrome: the role of small fiber damage. Clin Neurophysiol. 2026;187:2111881.

23. Grayston R, Czanner G, Elhadd K, et al. A systematic review and meta-analysis of the prevalence of small fiber pathology in fibromyalgia: implications for a new paradigm in fibromyalgia etiopathogenesis. Semin Arthritis Rheum. 2019;48(5):933–940.

24. Page MJ, McKenzie JE, Bossuyt PM, et al. The PRISMA 2020 statement: an updated guideline for reporting systematic reviews. BMJ. 2021;372:71.

25. Viechtbauer W. Conducting meta-analyses in R with the metafor package. J Stat Softw. 2010;36(3):1–48.

26. Ruggieri M, Gargano CD, Paparella G, et al. Neurofilament light chain in fibromyalgia: correlation with central and peripheral nervous system dysfunction. Eur J Pain. 2026;30(2):e70228.

27. Lauria G, Bakkers M, Schmitz C, et al. Intraepidermal nerve fiber density at the distal leg: a worldwide normative reference study. J Peripher Nerv Syst. 2010;15(3):202–7.

